# Awareness of the Importance of Genetic Counseling and Its Role in Preventing Genetic Disorders in Derna District

**DOI:** 10.64898/2026.01.27.25342786

**Authors:** Mohammed Al-Ghazali, Dina Idris Al-Mayar, Hadeel Osama Al-Fkhakhri, Hawa Mustafa Al-Hijazi

## Abstract

This study examined awareness, attitudes, and perceived barriers regarding genetic counseling among individuals in Derna District, focusing on its role in preventing genetic disorders. A descriptive cross-sectional design was employed, involving 278 participants aged 17 to 45 years, selected through stratified random sampling. Data were collected using structured questionnaires and analyzed with descriptive statistics via SPSS version 26.0. The findings revealed that while 65.5% of participants reported a high level of knowledge about genetic counseling, significant gaps remain, with 34.5% indicating low knowledge. Most participants demonstrated positive attitudes: 90.6% believed genetic counseling is important for preventing genetic disorders, and 90.3% expressed willingness to undergo counseling if recommended by a physician. However, perceived barriers such as fear of results (39.9%) and lack of awareness (30.9%) were reported. The study highlights the need for targeted educational initiatives and policy measures to promote genetic counseling services and address identified barriers. The findings provide valuable guidance for public health programs aiming to enhance the utilization of genetic counseling in the region.

## 1. Introduction

### 1.1 Background of the Study

Genetic counseling is an essential component of healthcare that provides individuals and families with information and guidance regarding inherited genetic conditions. It helps people understand their risk of inheriting or passing on genetic disorders, and it plays a key role in decision-making about family planning, medical care, and lifestyle choices. With advancements in genetic science, the importance of genetic counseling has grown, particularly in the prevention of genetic disorders.

In many regions, including the Derna District in Libya, awareness of genetic counseling is still limited, and individuals may not be fully informed about how genetic counseling can help prevent or manage genetic diseases. These disorders, which can affect a person’s health and quality of life, can often be prevented through early detection and proper genetic counseling. Understanding the role of genetic counseling can significantly improve public health outcomes by reducing the occurrence of preventable genetic conditions.

This study aims to assess the awareness of the importance of genetic counseling and its role in preventing genetic disorders in the Derna District. The findings of this research can help inform public health initiatives and educational programs to increase awareness and encourage the use of genetic counseling services.

### 1.2. The Role of Genetic Counseling: Key Reasons to Consult a Specialist

**Figure No. 1.**
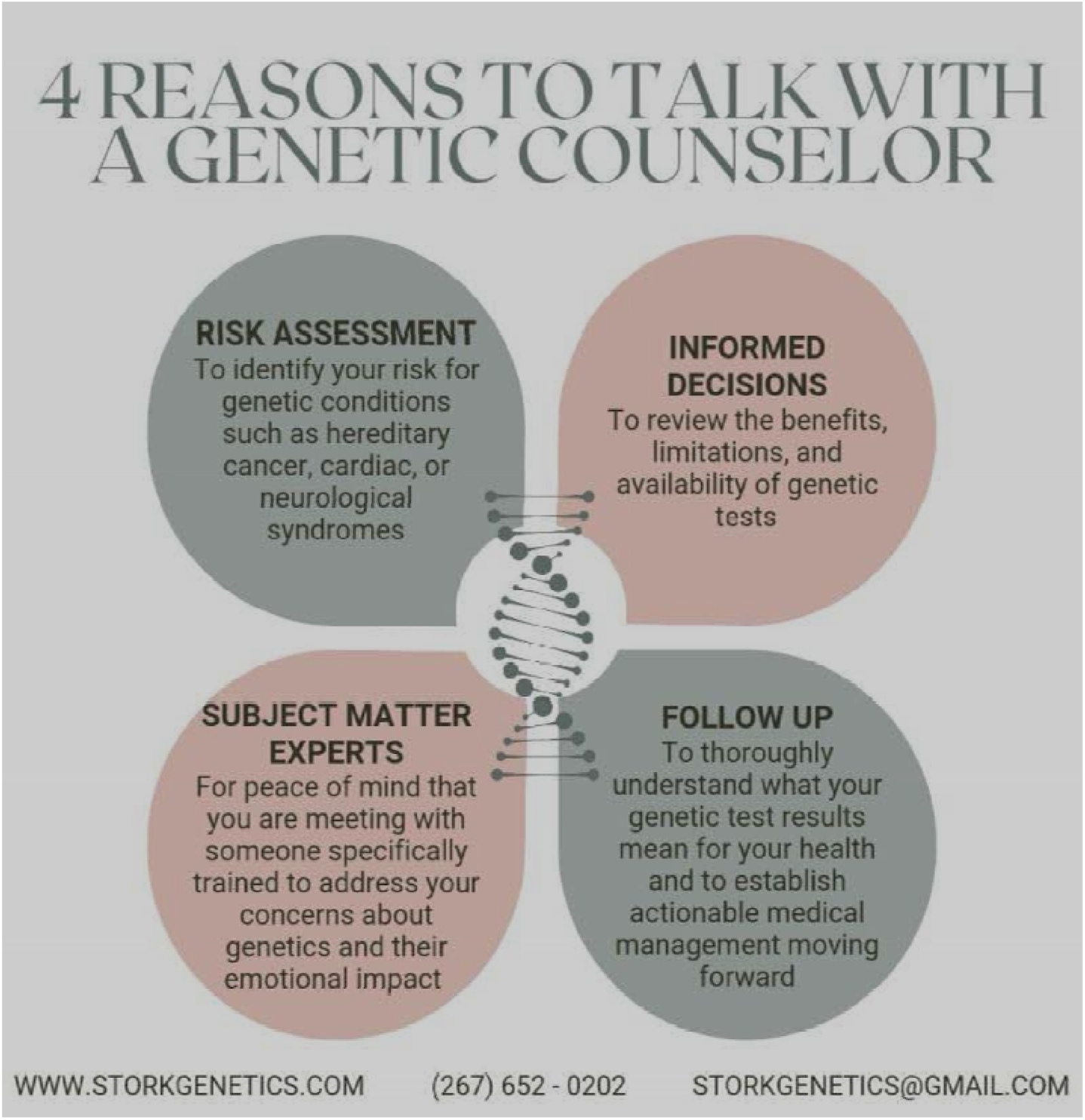
Outlines four primary reasons to consult a genetic counselor, emphasizing the importance of genetic counseling in managing and preventing genetic-related health concerns.

1. **Risk Assessment:**

- *Genetic counseling helps identify individual or familial risks for genetic conditions such as hereditary cancer, cardiac issues, or neurological syndromes*.
- *This ensures individuals have a clearer understanding of their predisposition to certain health conditions, enabling early intervention*.
2. **Informed Decisions:**

- *Counselors help individuals evaluate the benefits, limitations, and availability of genetic testing*.
- *This guidance empowers patients to make educated choices about whether to pursue genetic tests and how to interpret their implications*.
3. **Subject Matter Experts:**

- *Genetic counselors are professionals specifically trained to address the complexities of genetic health and provide emotional support*.
- *This expertise offers patients peace of mind, as their concerns are handled by qualified professionals*.
4. **Follow-Up:**

- *After genetic testing, counselors assist in interpreting results and planning appropriate medical management*.
- *This ensures individuals fully understand their results and have actionable steps to manage their health moving forward*.

### 1.3 Problem Statement

The lack of awareness regarding genetic counseling in Derna District presents a challenge in the prevention and management of genetic disorders. Many individuals are unaware of how genetic counseling can aid in early detection and the prevention of genetic diseases. This lack of awareness may contribute to the continued prevalence of preventable genetic disorders in the region.

### 1.4 Research Objectives

The primary objectives of this study are:

1. To assess the level of awareness of genetic counseling in Derna District.
2. To evaluate the perceived importance of genetic counseling in preventing genetic disorders.
3. To identify the barriers to accessing genetic counseling services in the region.
4. To provide recommendations for improving awareness and access to genetic counseling in Derna District.

### 1.5 Research Questions

1. What is the level of awareness of genetic counseling among the residents of Derna District
2. What are the attitudes of the population towards genetic counseling and its role in preventing genetic disorders
3. What are the main challenges or barriers to accessing genetic counseling services in the Derna District

### 1.6. Significance of the Study

This study is significant because it aims to provide insights into the awareness and attitudes of the public towards genetic counseling, which is crucial for preventing genetic disorders. By understanding these factors, health authorities can design targeted awareness campaigns and improve access to genetic counseling services, ultimately reducing the prevalence of genetic disorders in the region.

## 2. Literature Review

### 2.1 Genetic Counseling: Definition and Purpose

Genetic counseling is a process in which a trained healthcare professional provides individuals or families with information about genetic conditions and how they may affect them. The goal of genetic counseling is to help individuals understand genetic risk factors, make informed decisions, and manage the emotional aspects of genetic conditions (1).

| Study | Country | Key Findings | Methodology | Significance | Reference |
| --- | --- | --- | --- | --- | --- |
| Abdallah NA, et al. (2014) | Egypt | Found that 30% of participants were aware of genetic counseling; barriers included lack of information and cultural stigma. | Cross-sectional survey with 500 participants. | Highlighted the need for public health campaigns to raise awareness. | (7) |
| Al-Saleh M, et al. (2016) | Libya | Identified a severe shortage of genetic counselors and limited awareness among the public. | Interviews with 20 healthcare professionals. | Recommended government investment in genetic counseling training programs. | (8) |
| Trotter MJ, et al. (2016) | Saudi Arabia | Cultural stigma was a significant barrier to seeking genetic counseling services. | Mixed-methods: surveys and focus groups with 300 participants. | Suggested incorporating genetic counseling education into schools. | (10) |
| Mukhtar S, et al. (2018) | UAE | 45% of respondents had basic knowledge of genetic counseling, but only 10% had used the services. | Descriptive study with a questionnaire distributed to 1,000 participants. | Identified the potential of tele-genetic counseling to improve access. | (15) |
| Crozier R, et al. (2015) | United States | High awareness of genetic counseling (70%) due to better healthcare infrastructure and insurance coverage. | National health survey with 10,000 participants. | Emphasized the role of accessible services in raising awareness. | (12) |
| Hodgson S, et al. (2014) | United Kingdom | Awareness varied based on socioeconomic status; higher awareness in urban areas. | Stratified random sampling of 2,000 households. | Advocated for targeted awareness campaigns in rural areas. | (16) |
| Lemoine M, et al. (2017) | Tunisia | Limited awareness among women; cultural factors played a major role in underutilization. | Qualitative interviews with 50 women. | Recommended involving community leaders to break cultural barriers. | (14) |

In many cases, genetic counseling can help prevent the occurrence of genetic disorders by identifying individuals at risk and suggesting preventive measures. Genetic counselors use family history and genetic testing to assess risks and provide personalized advice. Counseling may include discussing reproductive options, lifestyle changes, or strategies for managing or preventing diseases (2).

### 2.2. Previous Studies on Awareness of Genetic Counseling in Different Countries

### 2.3 The Role of Genetic Counseling in Preventing Genetic Disorders

The role of genetic counseling in preventing genetic disorders cannot be overstated. It helps individuals and families make informed decisions based on genetic information, which can lead to better health outcomes. Studies have shown that individuals who receive genetic counseling are more likely to take preventive actions, such as undergoing genetic testing or making lifestyle adjustments, to reduce their risk of developing a genetic disorder (3, 4).

Research suggests that genetic counseling can be particularly valuable in the prevention of inherited conditions, such as cystic fibrosis, sickle cell anemia, and Tay-Sachs disease (5). For example, genetic counseling can help identify carriers of recessive genetic disorders, allowing for informed decisions about family planning, including the option of prenatal testing or considering the use of assisted reproductive technologies (6).

### 2.4 Awareness and Accessibility of Genetic Counseling

Awareness of genetic counseling varies widely across different regions. In developed countries, genetic counseling services are more accessible, and there is generally a higher level of awareness about their importance. However, in many developing countries, including Libya, awareness and access to these services are limited. A study conducted in Egypt found that only a small percentage of the population was aware of genetic counseling and its role in preventing genetic disorders (7).

In Libya, particularly in regions like Derna District, the situation is similar. While healthcare infrastructure in larger cities may be better equipped, rural areas often lack the necessary resources and public health education to raise awareness about genetic counseling. This lack of awareness may result in a lower demand for genetic counseling services, contributing to the persistence of genetic disorders in the population (8).

### 2.5 Barriers to Accessing Genetic Counseling

Several factors contribute to the underutilization of genetic counseling services. One significant barrier is the lack of trained genetic counselors. In many regions, there is a shortage of professionals in the field, which makes it difficult for individuals to access counseling services (9). Additionally, cultural factors, including stigmas around genetic conditions, may discourage people from seeking genetic counseling (10).

Economic factors also play a role. The cost of genetic testing and counseling services can be prohibitive, especially in low-income regions. In some cases, individuals may not be able to afford these services, even if they are aware of their availability (11).

### 2.6. Importance of Genetic Counseling in Public Health

Genetic counseling is essential for public health because it contributes to the prevention and management of genetic disorders, thereby reducing the social and economic burden of these conditions. By increasing awareness of genetic counseling, individuals are empowered to make decisions that can improve their quality of life and reduce the transmission of genetic disorders to future generations (12, 13).

Increasing the accessibility and awareness of genetic counseling services can lead to a healthier population and lower the incidence of preventable genetic conditions. Public health programs aimed at increasing awareness of genetic counseling can be a powerful tool in addressing the burden of genetic disorders in regions with limited access to healthcare resources (14, 15).

### 2.7. Study Benefits and Different

1. **Focus on Public Knowledge** Unlike studies that emphasize healthcare provider perspectives or service accessibility, this study directly evaluates public knowledge about genetic counseling in the **Derna District**, providing an understanding of the population’s awareness and misconceptions.
2. **Localized Context** Most previous studies target entire nations or large urban centers. This study focuses specifically on the Derna District, providing detailed insights into the knowledge gaps within this localized population, particularly in rural areas.
3. **Barriers and Misconceptions** This study not only assesses general awareness but also investigates the barriers (e.g., cultural beliefs, misinformation) that prevent people from understanding or utilizing genetic counseling services effectively.
4. **Comparative Insights** By comparing findings with similar studies in other countries (e.g., UAE, Egypt, Saudi Arabia), the study offers a global perspective while highlighting regional differences specific to Derna District.
5. **Actionable Recommendations** The study’s results will inform the design of public health interventions tailored to the cultural and social norms of Derna District, ensuring effective educational campaigns to improve public knowledge and acceptance of genetic counseling.
6. **Focus on Preventive Knowledge** Many previous studies focused on attitudes or barriers to service use. This study emphasizes the population’s *knowledge of the preventive role* of genetic counseling, which is critical for reducing genetic disorders.

## 3. Methodology

### 3.1 Research Design

This study will adopt a descriptive cross-sectional design to assess the awareness and attitudes towards genetic counseling in Derna District. A cross-sectional design is suitable for this research as it allows for the collection of data at a single point in time, providing a snapshot of public knowledge and perceptions of genetic counseling (16).

**Figure No. 2.**
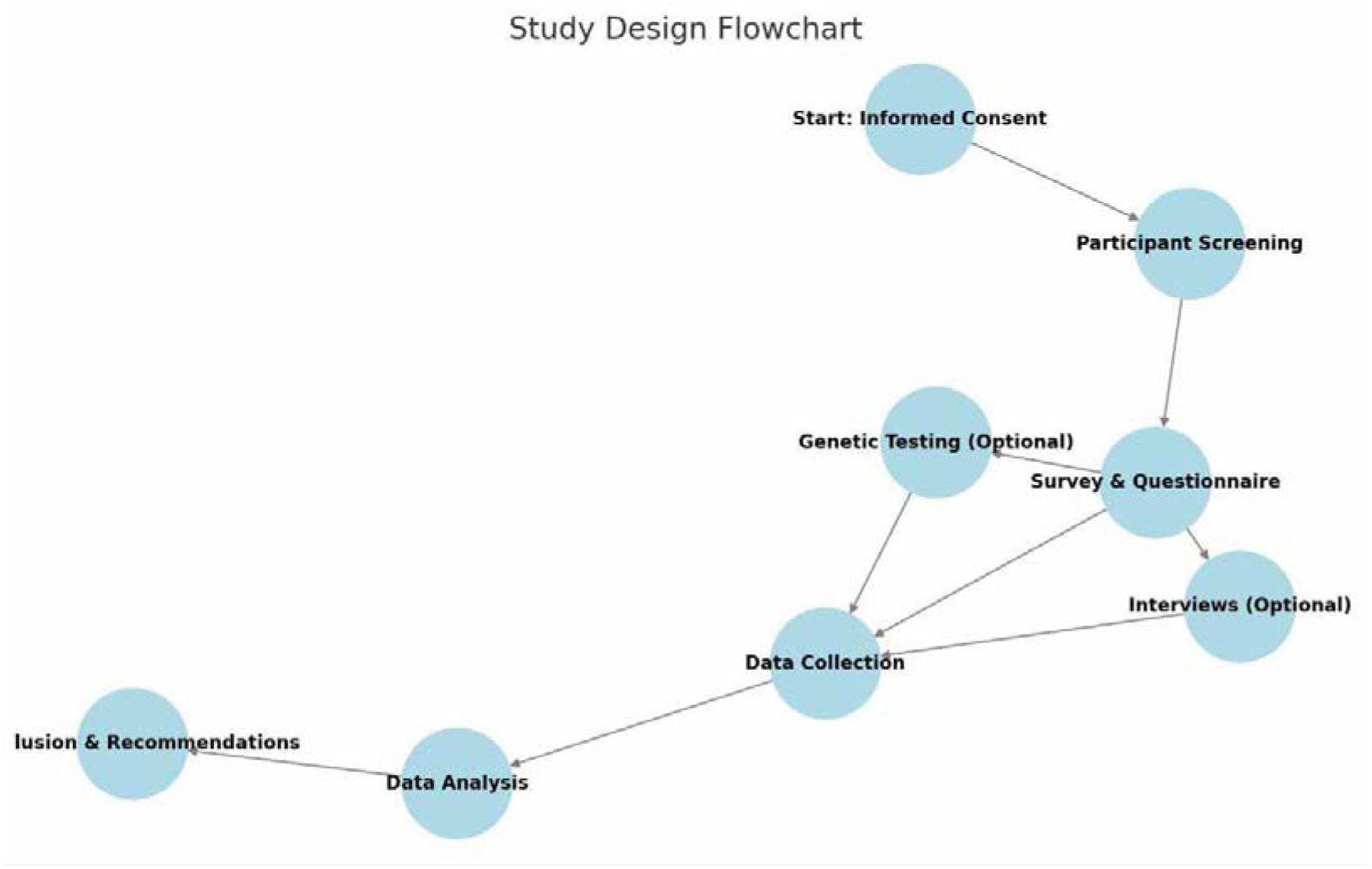
Design to assess the awareness and attitudes towards genetic counseling in Derna District.

### 3.2 Study Population

The study will target residents of Derna District, a number of 300 individual will be randomly selected targeting those recently married and bachelor couples, with a focus on both urban and rural areas. The population will include men and women of reproductive age, as they are most likely to seek genetic counseling for family planning and the prevention of genetic disorders. The sample will be selected using stratified random sampling to ensure a representative sample of the population from different demographic groups (17).

### 3.3 Data Collection

Data will be collected through structured interviews and questionnaires to a number of 300 individual randomly selected targeting those recently married and bachelor couples. The questionnaire will include questions related to the participants’ knowledge of genetic counseling, their attitudes towards its importance, and their experiences with genetic counseling services. Additionally, demographic information such as age, gender, education level, and employment status will be gathered to examine potential differences in awareness based on these factors.

### 3.4 Data Analysis

The collected data will be analyzed using descriptive statistics, including frequencies, percentages, and averages. This will allow for an overview of the general level of awareness and the factors that influence people’s attitudes towards genetic counseling. Inferential statistics, such as chi-square tests, may also be used to assess the relationship between demographic factors and the awareness of genetic counseling.

### 3.5 Ethical Considerations

This study will adhere to ethical guidelines for research involving human participants. Participants will be informed about the purpose of the study and their rights, including the right to confidentiality and the right to withdraw from the study at any time without penalty. Consent will be obtained from all participants before data collection begins.

## 4. Statistical Analysis

The collected data were analyzed using the Statistical Package for Social Sciences (SPSS) version 26.0 software. Descriptive data analysis were used. Mean and standard deviation.

### 4.1. Results

The study sample comprised 278 individuals, with ages ranging from 17 to 45 years and a mean age of 24.65 years (SD = 4.70). The majority of participants were younger than 25 years, accounting for 67.3% of the sample (n = 187). Those aged 25 to 35 years constituted 29.9% (n = 83), while a small minority, 2.9% (n = 8), were older than 35 years.

The majority of participants were female, comprising 222 individuals (79.9%), while male participants accounted for only 56 individuals (20.1%) (Table 1).

**Figure No. 3.**
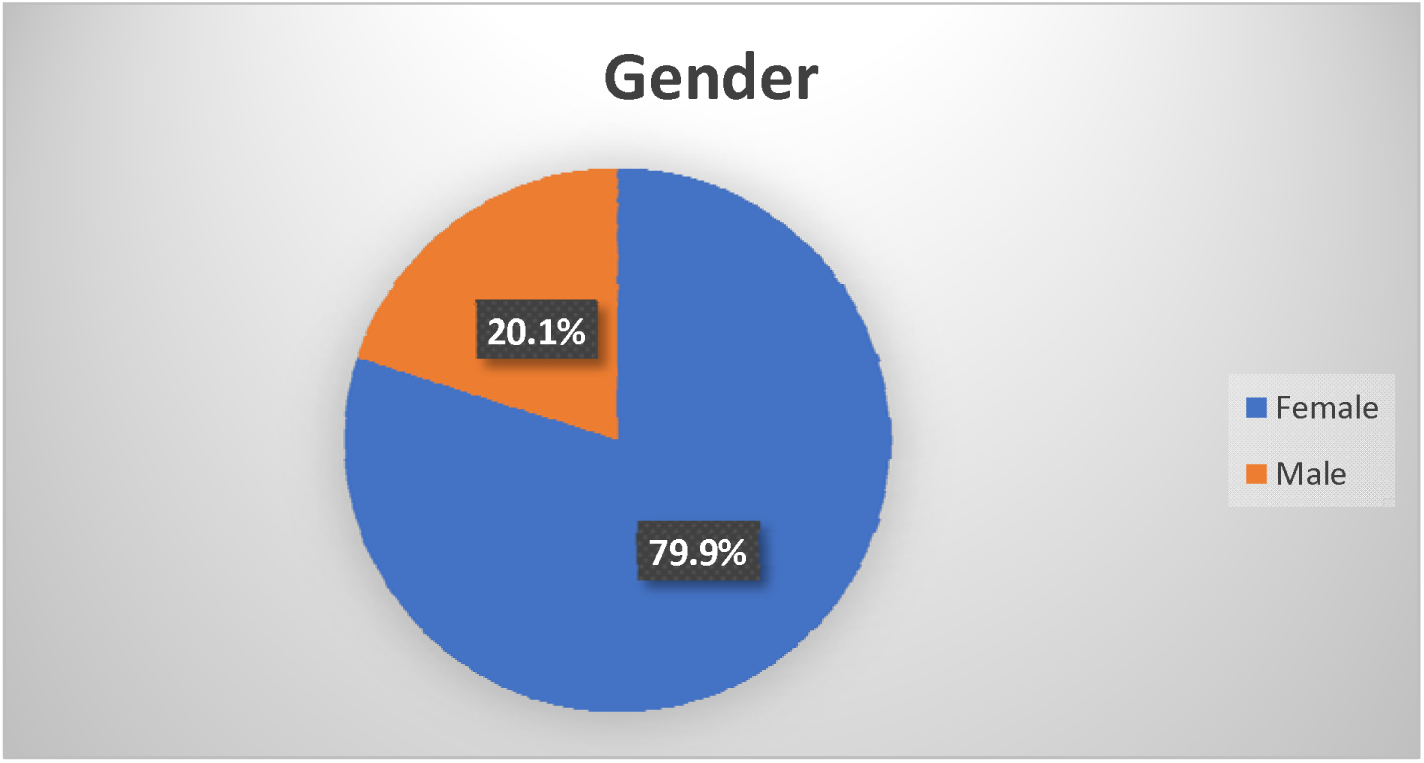
Pie Chart Showing Demographic Characteristics of Participants.

**Figure No. 4.**
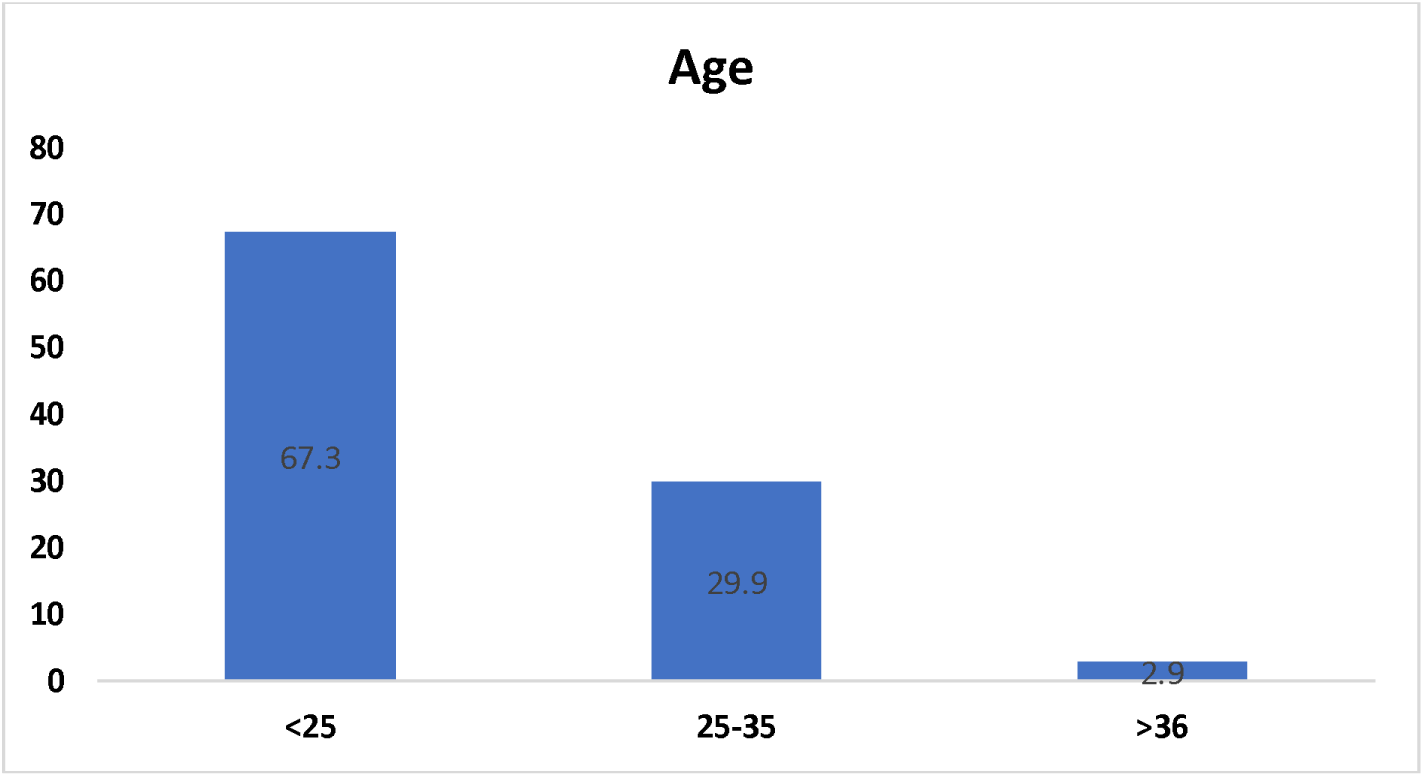
Chart Showing Age of Participants.

**Figure No. 5.**
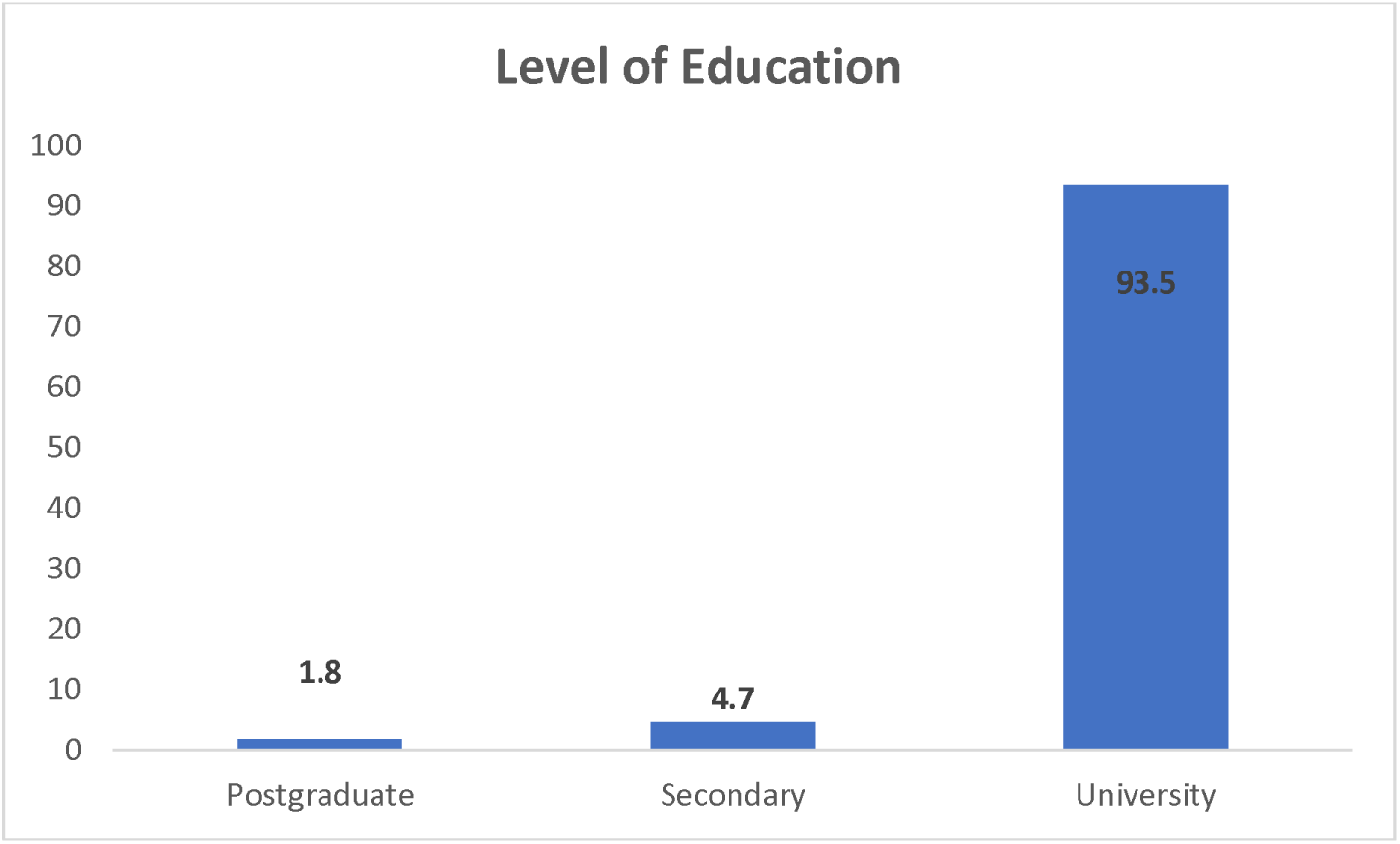
Column Chart Showing Level of Education of Participants.

**Table 1:**
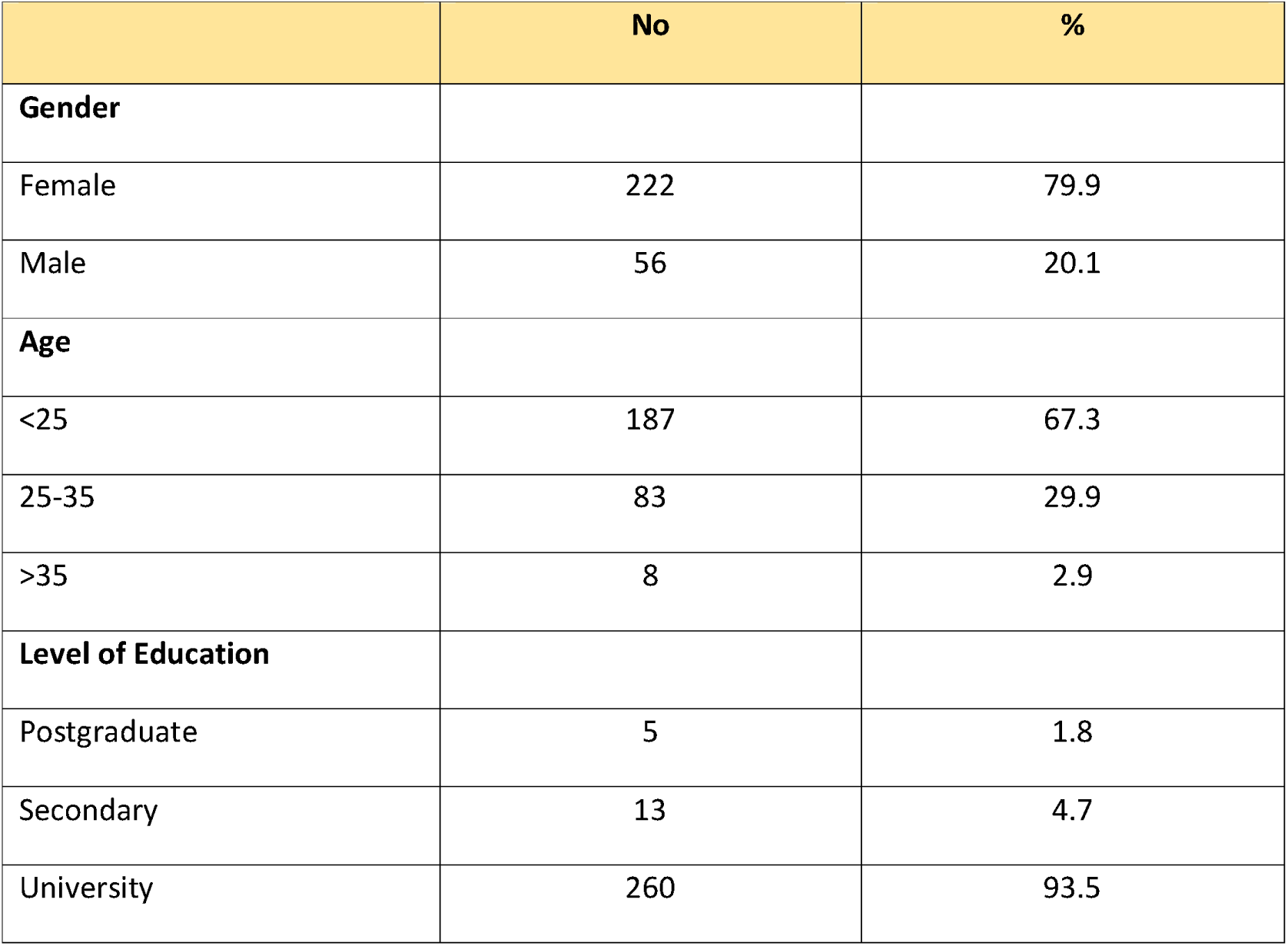
Demographic Characteristics of Participants.

Table 2 presents data on the participants’ family history of genetic disorders. Out of the total sample, 71 individuals (25.5%) reported having a family history of genetic disorders, while the remaining 207 participants (74.5%) indicated no such history.

**Figure No. 6.**
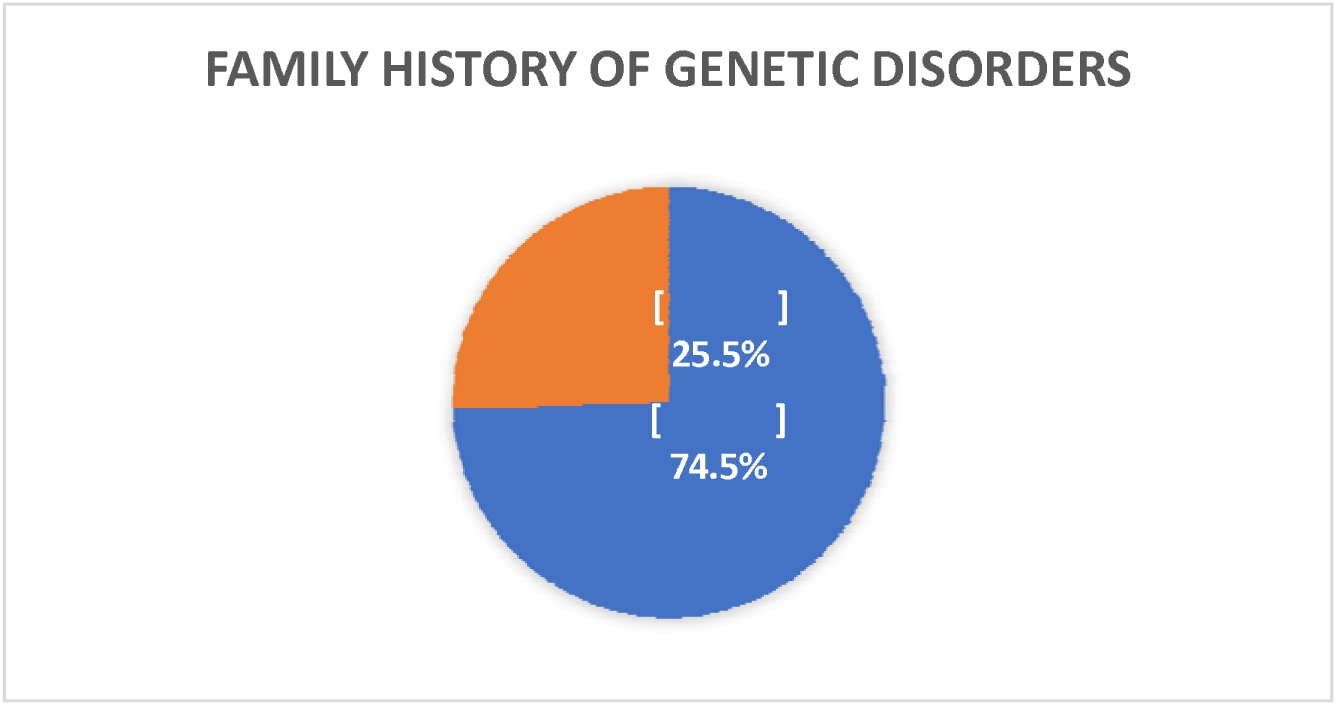
Pie Chart Showing Family History of Genetic Disorders among the Participants.

**Table 2:** Family history of genetic disorders.

|  | No | % |
| --- | --- | --- |
| No | 207 | 74.5 |
| Yes | 71 | 25.5 |

Among the 278 respondents, 58.3% (n = 162) reported having heard of genetic counseling, while 41.7% (n = 116) had not. When asked to evaluate their knowledge, 24.5% rated it as good, 23.7% as moderate, 17.3% as excellent, 13.3% as poor, and 21.2%indicated having no knowledge at all (Table 3).

**Table 3:** Participants’ Awareness of Genetic Counseling.

|  | No | % |
| --- | --- | --- |
| **Have you ever heard of genetic counseling?** |  |  |
| No | 116 | 58.3 |
| Yes | 162 | 41.7 |
| **How would you rate your knowledge of genetic counseling?** |  |  |
| Excellent | 48 | 17.3 |
| Moderate | 66 | 23.7 |
| Good | 68 | 24.5 |
| Poor | 37 | 13.3 |
| None | 59 | 21.2 |

The study indicated that (65.5%) of participants reported having a high level of knowledge about Genetic Counseling, while (34.5%) of participants reported having a low level of knowledge in this field (Figure 1).

**Figure No. 6.**
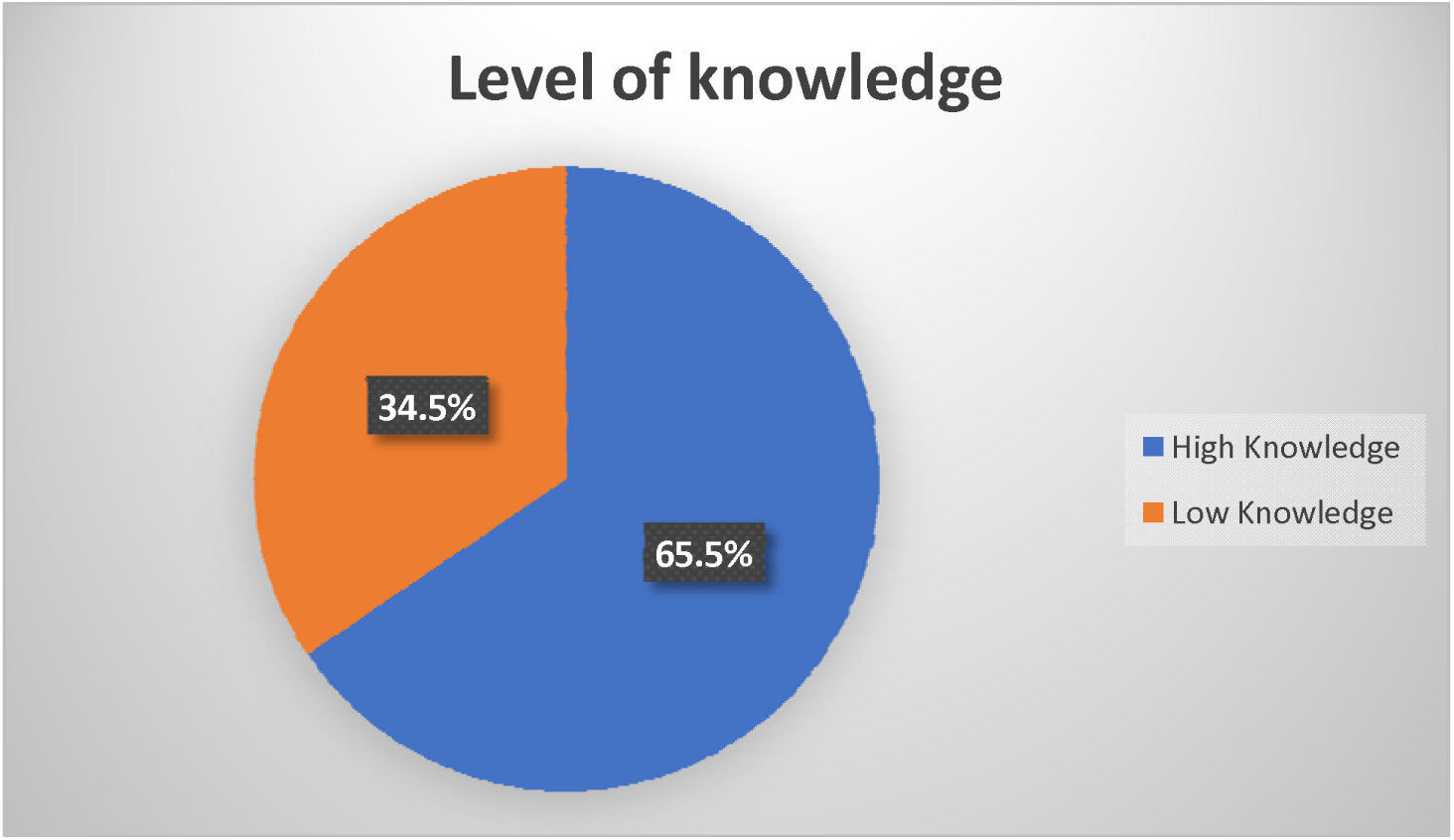
Pie Chart Showing Level of Knowledge among the participants.

Regarding attitudes toward the importance of genetic counseling, an overwhelming 90.3% (n = 252) believed that it plays a key role in preventing genetic disorders, compared to only 9.4% (n = 26) who did not share this view. Similarly, when asked whether they would consider undergoing genetic counseling if recommended by a doctor, 90.3% (n = 251) responded affirmatively, while 9.7% (n = 27) declined (Table 4).

**Table 4:** Participants’ Attitudes Toward Genetic Counseling.

|  | No | % |
| --- | --- | --- |
| **Do you believe genetic counseling is important in preventing genetic disorders?** |  |  |
| No | 26 | 9.4 |
| Yes | 252 | 90.6 |
| **Would you consider undergoing genetic counseling if recommended by a doctor?** |  |  |
| No | 27 | 9.7 |
| Yes | 251 | 90.3 |
| **Do you think genetic counseling should be mandatory before marriage?** |  |  |
| No | 58 | 20.9 |
| Yes | 220 | 79.1 |

When asked whether genetic counseling should be mandatory before marriage, 79.1% (n = 220) agreed, and 20.9% (n = 58) disagreed.

The study indicated that (86.7%) of participants reported having a high good attitudes about Genetic Counseling, while (13.3%) of participants reported bad attitudes (Figure 2).

**Figure No. 7.**
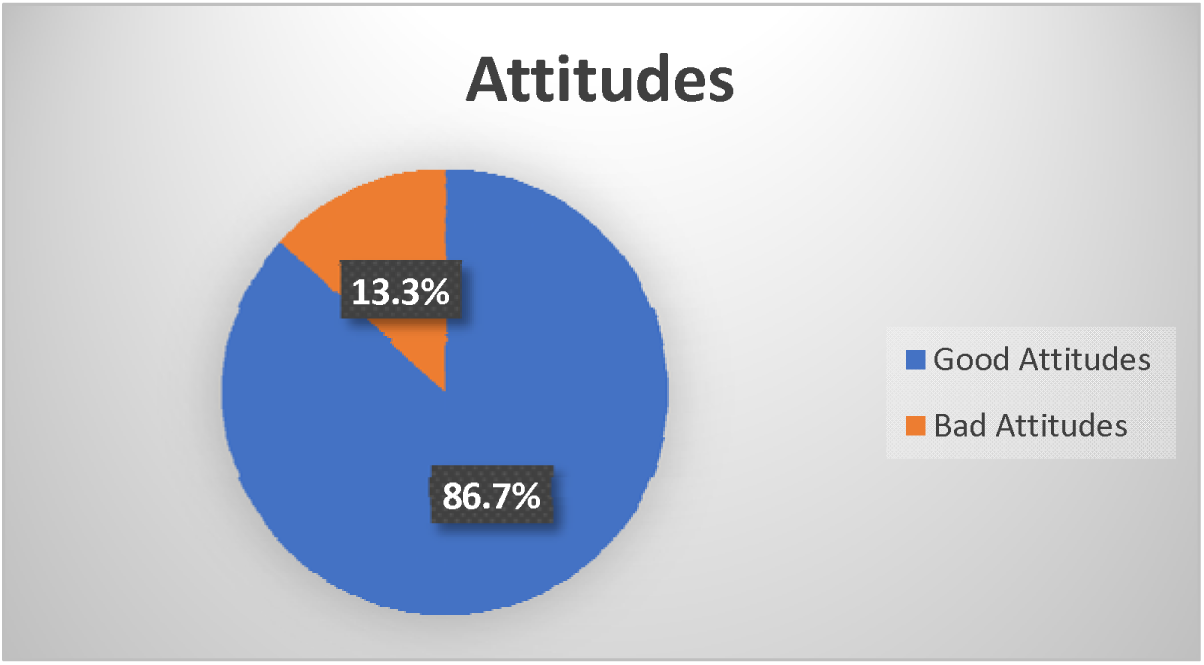
Pie Chart Showing the Attitudes among the participants.

**Table 5:** Perceived Barriers to Genetic Counseling.

|  | No | % |
| --- | --- | --- |
| **What do you think are the barriers to genetic counseling?** |  |  |
| Cultural beliefs | 50 | 18.0 |
| Fear of results | 111 | 39.9 |
| High cost | 56 | 20.1 |
| Lack of awareness | 86 | 30.9 |

Participants were also asked to identify perceived barriers to genetic counseling. The most commonly cited barrier was fear of results 39.9 %(n = 111), followed by lack of awareness 30.9%(n = 86), high cost 20.1%(n = 56), and cultural beliefs 18.0 %(n = 50) (Table 4).

### 4.2. Discussion

This study aimed to assess awareness, attitudes, and perceived barriers regarding genetic counseling among individuals aged 17 to 45 years. The findings revealed that while a majority of participants (65.5%) reported a high level of knowledge about genetic counseling, a significant proportion (34.5%) still exhibited low awareness. This suggests that although genetic counseling is increasingly recognized, gaps in knowledge remain within the community.

The results showed that most participants (90.6%) believed that genetic counseling is important in preventing genetic disorders, and a similar proportion (90.3%) indicated willingness to undergo counseling if recommended by a physician. These findings align with previous studies that have highlighted the positive attitude of populations toward genetic counseling when they are informed about its benefits. Furthermore, the fact that 79.1% of respondents supported mandatory genetic counseling before marriage reflects a strong endorsement for preventive health measures at a community level.

Despite the generally positive attitudes, barriers such as fear of results (39.9%) and lack of awareness (30.9%) were prominently reported. This highlights ongoing challenges in promoting genetic services. Cultural beliefs and high costs, though reported less frequently, still represent significant concerns that may hinder access to counseling services. These findings emphasize the need for targeted educational campaigns and subsidized services to address misconceptions and financial barriers.

## 5. Conclusion

In conclusion, this study demonstrated that while there is generally good awareness and highly positive attitudes towards genetic counseling among participants, important barriers persist. Fear of potential results and lack of awareness are primary obstacles that could limit the utilization of genetic counseling services. Efforts to improve public understanding and reduce anxiety associated with genetic testing are critical to enhancing acceptance and uptake of these services. The high level of support for mandatory premarital counseling indicates community readiness for policy initiatives in this area.

### 5.1. Limitations

This study had several limitations. First, the sample predominantly comprised young adults and females, which may limit the generalizability of the findings to the broader population. Second, the cross-sectional design captures perceptions at a single point in time and does not account for changes over time. Finally, the reliance on self-reported data may introduce response bias, as participants might have overestimated their knowledge or provided socially desirable responses regarding their attitudes.

### 5.2. Recommendations

Based on the findings, the following recommendations are proposed:

1. Public awareness campaigns should be implemented to improve understanding of genetic counseling and address common fears and misconceptions.
2. Healthcare providers should be trained to effectively communicate the benefits of genetic counseling and alleviate patients’ anxiety about results.
3. Subsidization or insurance coverage for genetic counseling services should be considered to reduce the barrier of high cost.
4. Policy initiatives, including the introduction of mandatory premarital genetic counseling, could be explored, given the high level of community support.
5. Further research should be conducted on diverse populations, including rural areas and different age groups, to obtain more representative data.

## Data Availability

https://www.researchgate.net/profile/Mohammed-Alghazali-2/stats

## 5.3. Declarations

### 5.3.1. Ethics Approval and Consent to Participate

Ethical approval for this study was **waived** by the **Scientific Research Ethics Committee, Faculty of Medical Technology – Derna, University of Derna, Libya**, as the study involved an anonymous voluntary questionnaire survey, did not include any personal identifiers, and posed no risk to participants.

Informed consent was obtained from all participants prior to participation, and confidentiality and anonymity of all collected data were strictly maintained in accordance with institutional research ethics guidelines.

#### 5.3.1.1. Consent for Publication

Not applicable.

#### 5.3.1.2. Availability of Data and Materials

The datasets used and/or analyzed during the current study are available from the corresponding author on reasonable request.

#### 5.3.1.4. Funding

This research received no specific grant from any funding agency in the public, commercial, or not-for-profit sectors.

#### 5.3.1.5. Authors’ Contributions

Mohammed Al-Ghazali, B.Sc., M.Sc. Med, conceived and designed the study, supervised data collection, performed the statistical analysis, and prepared the final manuscript.

All student co-authors — Mrs. Dina Idris Al-Mayar, B.Sc. Med. Tech. (Genetic Eng.), Ms. Hadeel Osama Al-Fkhakhri, B.Sc. Med. Tech. (Genetic Eng.), and Ms. Hawa Mustafa Al-Hijazi, B.Sc. Med. Tech. (Genetic Eng.) — contributed to data collection and questionnaire administration, and approved the final manuscript.

